# Genetic susceptibility to pneumonia: A GWAS meta-analysis between UK Biobank and FinnGen

**DOI:** 10.1101/2020.06.22.20103556

**Authors:** Adrian I. Campos, Pik Fang Kho, Karla X. Vazquez-Prada, Luis M. García-Marín, Nicholas G. Martin, Gabriel Cuéllar-Partida, Miguel E. Rentería

**Author notes:** Correspondence: Adrian I. Campos. 23andMe Inc, Sunnyvale, CA, USA.

## Abstract

**Rationale:** Pneumonia is a respiratory condition with complex aetiology. Host genetic variation is thought to contribute to individual differences in susceptibility and symptom manifestation.

**Methods:** We analysed pneumonia data from the UK Biobank (14,780 cases and 439,096 controls) and FinnGen (9,980 cases and 86,519 controls). We perform genome-wide association study (GWAS) meta-analysis, gene-based test, colocalisation, genetic correlation, latent causal variable and polygenic prediction in an independent Australian sample (N=5,595) to draw insights into the genetic aetiology of pneumonia risk.

**Results:** We identify two independent loci on chromosome 15 (lead SNPs rs2009746 and rs76474922) to be associated with pneumonia(p<5e-8). Gene-based tests revealed eighteen genes in chromosomes 15,16 and 9, including *IL127, PBX3, APOBR* and smoking related genes *CHRNA3/5*, associated with pneumonia. Evidence of *HYKK* and *PBX3* involvement in pneumonia risk was supported by eQTL colocalisation analysis. We observed genetic correlations between pneumonia and cardiorespiratory, psychiatric and inflammatory related traits. Latent causal variable analysis suggests a strong genetic causal relationship cardiovascular health phenotypes and pneumonia risk. Polygenic risk scores (PRS) for pneumonia significantly predicted self-reported pneumonia history in an independent Australian sample, albeit with a small effect size (OR=1.11 95%CI=[1.04-1.19], p<0.05). Sensitivity analyses suggested the associations in chromosome 15 are mediated by smoking history, but the association of genes in chromosome 16 and 9, and polygenic prediction were robust to adjustment for smoking.

**Conclusions:** Altogether, our results highlight common genetic variants, genes and potential pathways that contribute to individual differences in susceptibility to pneumonia, and advance our understanding of the genetic factors underlying heterogeneity in respiratory medical outcomes.

## INTRODUCTION

Pneumonia is an inflammatory condition of the lungs that usually stems from an infection. It is characterised by alveolar filling with fluid, microorganisms and immune response cells, preventing the lungs from working properly.^1^ Diagnosis is confirmed with chest radiography showing abnormalities, and other pieces of evidence such as laboratory tests identifying the causal pathogen and increases in antibody count^2^. Pneumonia is associated with increased morbidity and mortality;^3^ in fact, mortality estimates range between five and 14% for hospitalised patients. Risk factors for pneumonia include smoking,^4^ alcoholism,^5^ heart disease, and advanced age^6^. Furthermore, mortality amongst pneumonia cases is associated with factors such as hypertension and smoking.^7^ Nonetheless, individuals considered ‘*at low risk*’ of pneumonia can still develop the condition, which highlights its complexity and clinical heterogeneity.

Since the emergence of the 2020 COVID-19 pandemic, there has been an increase in pneumonia incidence and mortality.^8^ Its relatively high infectivity and mortality even among *low-risk* groups calls for the investigation of genetic mechanisms underlying pathogenesis and prognosis. A recent study on 2633 British twins (728 complete pairs, 537 monozygotic and 191 dizygotic, 86.9% female) investigated the susceptibility to infection by SARS-CoV-2.^9^ The researchers used a symptom-based algorithm to predict true infection in participants tested for SARS-CoV-2 and estimated heritability for symptoms including fever = 0.41 (95% CI 0.12-0.70); anosmia 0.47 (0.27-0.67) and delirium 0.49 (0.24-0.75). Overall predicted heritability of COVID-19 status was 0.50 (0.29-0.70), suggesting that symptomatic infection with SARS-CoV-2 is under host genetic influence to some extent, and reflecting inter-individual variation in the host immune response. Thus, host-specific genetic susceptibility is an emerging area of research interest^10^ as it could facilitate the systematic stratification of patients by genetic risk and aid in the design of more efficient treatments.^11^

In fact, evidence from other infectious diseases points to an important role for host genetics in influencing the development of symptomatic infection.^12^ Twin studies have shown higher concordance rates of tuberculosis, leprosy, poliomyelitis and hepatitis B in identical versus non-identical twins, suggesting a genetic component in susceptibility to these infectious diseases.^12^ Moreover, clinical trials for drugs targeting genes with evidence of disease association are more likely to lead to useful therapies.^13,14^ Thus, identification of genes and pathways that confer increased susceptibility to pneumonia could reveal new therapeutic targets and inform the design of prevention and treatment strategies.

Here, we report a GWAS meta-analysis of pneumonia history in adults using data from two large datasets, the UK Biobank and FinnGen. We identify genetic variants and genes associated with pneumonia risk, an essential step for understanding inter-individual differences in susceptibility. We characterise the genetic aetiology of pneumonia by assessing its genetic correlations and genetic evidence for causality against ∼1,500 traits with publicly available GWAS data. Finally, we demonstrate the external validity of our findings by performing polygenic prediction of self-reported pneumonia in an independent Australian sample.

## METHODS

### Samples and phenotypic information

For this study, we meta-analysed GWAS for pneumonia in two independent samples: the UK Biobank and FinnGen. For UK Biobank, we conducted a GWAS of pneumonia using individual-level genetic and phenotypic data from the UK Biobank. International Classification of Diseases (ICD10) codes are used to store information on participants’ health conditions. Raw ICD10 data were extracted from the UK Biobank under Application Number 25331. In this study, we excluded participants of non-European ancestry to avoid potential genetic associations emerging from population stratification. Participants with a history of pneumonia were defined as those presenting any ICD10 code related to infectious pneumonia (N=14,780) (see **Supplementary Table 1**). For FinnGen, we leveraged publicly available summary statistics on the phenotype *ICD10-J10 pneumonia* which comprised 9,980 cases and 86,519 controls. Information on sample phenotyping, genotyping and GWAS in the FinnGen sample is available elsewhere.^15^

### Pneumonia GWAS in the UK Biobank

GWAS was performed using BOLT-LMM, which implements a linear mixed model association analysis and fits a genetic relationship matrix as a random effect to account for cryptic relatedness and population stratification. Age, sex, genotyping array and the first 20 genetic principal components were adjusted for in the analysis. We used a stringent quality control procedure corresponding to minor allele frequency (MAF?≥?0.01) and imputation quality (INFO≥0.60).

### GWAS meta-analysis

A z-score meta analysis of pneumonia summary statistics was conducted between the UK Biobank and Finngen samples using METAL v(2011-03-25). The final meta-analysis comprised 24,760 cases and 525,615 controls. Only variants passing quality control in both cohorts were included in the meta-analysis. Furthermore, variants with inconsistent allele frequencies in both cohorts (difference >0.15) were removed. The final number of variants meta-analysed and included in this study was 7,831,927. Independent genetic signals were identified by clumping (r2<0.05, and 1Mb window) using CTG-VL (beta 0.1)^16^. A sensitivity analysis was performed by adjusting the GWAS results using multi-trait conditional and joint analysis (mtCOJO) to simultaneously adjust for two smoking phenotypes: smoking history and cigarettes per day.

### Gene-based analysis

Gene-based analysis was conducted on both the main and smoking adjusted GWAS using the “set-based association analysis for human complex traits” fastBAT method^17^ available on CTG-VL (https://genoma.io). fastBAT performs a set-based enrichment analysis based on the GWAS summary statistics while accounting for linkage disequilibrium (LD) between SNPs. We tested the association between 24,443 genes and pneumonia using this method. Statistical significance was defined using Benjamini-Hochberg False Discovery Rate (FDR) < 5% for multiple testing correction. Genes identified as statistically significant were further assessed for eQTL colocalisation with pneumonia.

### Colocalisation and eQTL

To assess the co-occurrence of signals in GWAS data and cis-expression quantitative trait loci (eQTL) data, we performed a summary-based colocalisation analysis. We integrated our GWAS data and cis-eQTL data from lung tissue and whole blood in GTEx V7. We used GWAS and eQTL summary statistics of SNPs within 1Mb window around each fastBAT-identified gene to estimate the posterior probability that GWAS signals co-occur with eQTL signals while accounting for LD structure. This method estimates the posterior probabilities for five different scenarios: no association with either trait (PP0), association with the disease only (PP1), association with gene expression only (PP2), associations with both traits but distinct SNPs (PP3) and associations with both traits in same SNPs (PP4). A threshold of PP4/(PP3+PP4) > 0.8 was considered as evidence for co-occurrence of GWAS signals and eQTL signals at the region of interest. Colocalisation analysis was performed using the *COLOC* package in R.

### Heritability and genetic correlations

We used LD-score regression (LDSC) to estimate the SNP-based heritability (h_SNP^2^_) for pneumonia on the liability scale assuming prevalence estimates of UK Biobank (3.3%) as both sample and population prevalence. Genetic correlations (rG) between pneumonia and 1,522 phenotypes were estimated using bivariate LDSC regression in CTG-VL based on a common set of HapMap3 variants. Benjamini-Hochberg FDR at 5% was used to assess statistical significance.

### Genetic Causal Proportion

To assess whether significant genetic correlations observed could be explained by an underlying causal relationship between traits, we used the Latent Causal Variable (LCV) method^18^ as implemented in CTG-VL. LCV uses GWAS summary statistics to estimate the genetic causal proportion (GCP) between two traits. The GCP’s absolute value ranges from 0 (no genetic causality) to 1 (full genetic causality). In our study, a high GCP value (GCP > 0.60) indicates that pneumonia is likely to affect the trait of interest. In contrast, a robust negative value (GCP < -0.60) provides evidence that the trait of interest is likely to affect pneumonia. For traits of interest (deep vein thrombosis, LDL and cholesterol) with significant evidence of a causal effect on pneumonia, generalised summary data-based Mendelian Randomisation (GSMR) was used as a secondary assessment of the existence of a causal relationship.

### Target sample and polygenic risk scoring

To assess the external validity of the GWAS, we performed polygenic based prediction on an independent target sample of 5,595 unrelated Australian Adults from the Australian Genetics of Depression Study (AGDS) with complete data.^19^ Pneumonia cases were identified through self-reported medical history in AGDS. PRS analysis was further adjusted for smoking by: i) additionally including smoking history as a covariate and ii) performing PRS calculation using the summary statistics adjusted for smoking history and cigarettes per day. Smoking history was assessed with the item: “*Have you smoked more than 100 cigarrettes in your lifetime*?”. We employed a recently developed method, SBayesR, to obtain the conditional effects of the studied variants, thus avoiding inflation arising from using correlated SNPs due to LD. Pneumonia polygenic risk scores (PRS) were calculated using PLINK 1·9. in the AGDS sample. Briefly, a PRS is calculated by multiplying the effect size of a given risk allele (obtained from the discovery GWAS summary statistics) by the imputed number of risk alleles (using dosage probabilities) present in each individual. Then, obtaining a weighted average across all loci. To assess the association between pneumonia PRS and self-reported pneumonia history in AGDS, we used a logistic regression model (python *statsmodels*). Pneumonia PRS was the predictive variable of interest, with age, sex and the first 20 genetic ancestry principal components included as covariates.

## RESULTS

### Prevalence of pneumonia and sample demographics

The prevalence of lifetime pneumonia in the UK Biobank was 3.3%. Sex was associated with pneumonia, where females were less likely to have experienced the condition (Female OR = 0.713 95%C.I.=[0.69-0.737]). Furthermore, participants with a history of pneumonia were on average older than controls (OR = 1.06 95%C.I.= [1.06-1.07]). Smoking history was also associated with an increased pneumonia risk (OR=1.74 95%C.I.=[1.68-1.68] see **Table 1**).

**Table 1.** GWAS UK Biobank sample composition.

| Table 1. GWAS UK Biobank sample composition |  |  |  |
| --- | --- | --- | --- |
|  | Cases | Controls | OR (95% C.I.) |
| Sample size | 14780 (3.3%) | 439096 (96.7%) | NA |
| Female N(%) | 6490 (44%) | 240059 (55%) | 0.713 (0.69-0.737) |
| Age mean(sd) | 60.4 (7.2) | 56.7 (8.0) | 1.06 (1.06-1.07) |
| Smoking history N(%) | 9143 (62%) | 198667 (45%) | 1.74 (1.68-1.68) |
| Data for participants of European ancestry included in the GWAS |  |  |  |

### Pneumonia GWAS

Our GWAS meta-analysis identified two independent genome-wide significant variants on 15q15.1 (index SNPs rs2009746 and rs76474922; p<5e-8; **Figure 1a**). The significant locus was located in a gene-rich region near *IREB2, CHRNA3/5* and *HYKK* (**Supplementary Figure 1**). In addition, eighteen independent loci showed suggestive association with pneumonia (**Table 2**). The amount of variance on the liability of pneumonia explained by this GWAS in the UK Biobank, also called the SNP heritability of the trait, the whole meta-analysis was estimated at 0.03 (s.e.=0.006) using LDSC regression. A sensitivity analysis using mtCOJO to adjust for smoking history and cigarettes per day revealed the hits on chromosome 15, but not other signals, to be mediated by smoking. A near genome-wide signal in chromosome 3, near the gene *SUCNR1*, became significant after conditioning on smoking phenotypes (**Figure 1b**) Notably, the genetic correlation between the unconditional and smoking conditional GWAS was high (rg=0.9371, S.E.= 0.015).

**Table 2.**
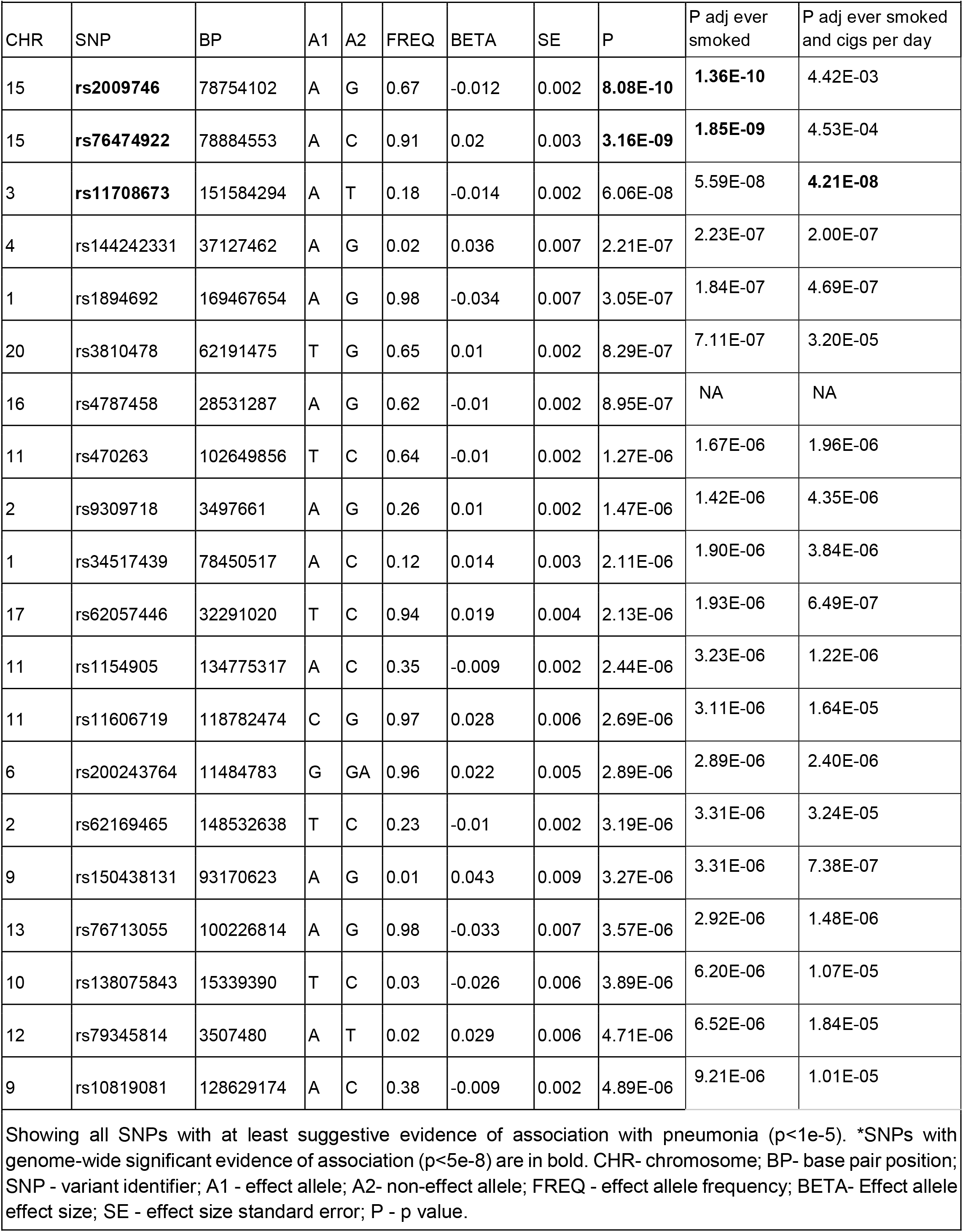
Pneumonia GWAS meta analysis and sensitivity results.

**Figure 1.**
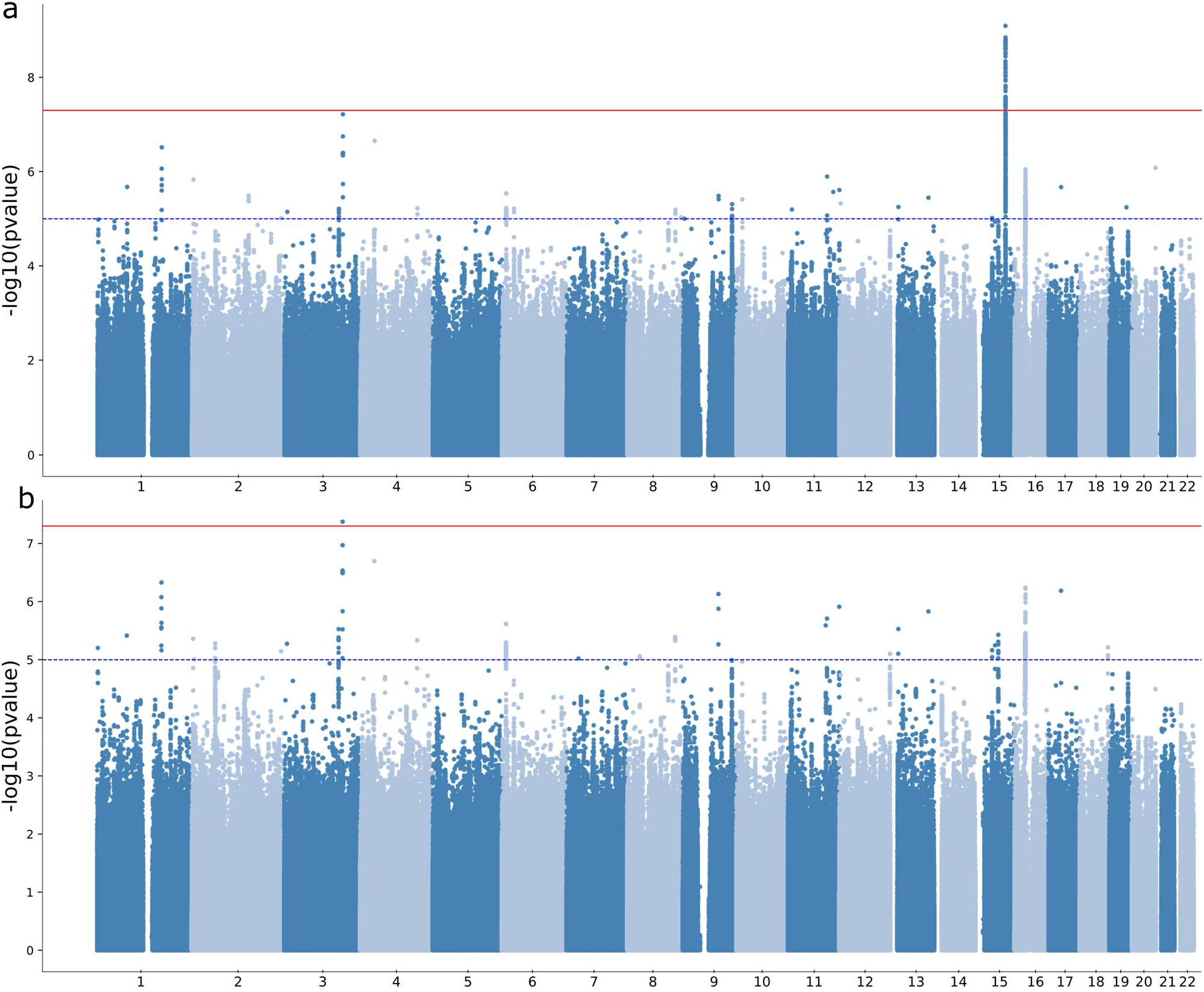
Pneumonia GWAS meta-analysis and gene based association tests. **a)** Manhattan plot shows the results of the genome-wide association study meta-analysis. Each dot represents a genetic variant. The x-axis is the genomic location ordered by chromosome. The y-axis represents the statistical evidence of the association (-log10 transformed p-value). The solid red and dashed blue lines represent the genome-wide and suggestive association significance thresholds. **b)** Manhattan plot shows the results of a sensitivity analysis using mtCOJO to condition on smoking history and cigarettes per day. Note the hit on chromosome 15 is no longer significant after this adjustment, while other signals remain largely unchanged.

### Gene-based analysis and colocalisation

We performed gene-based association testing followed by colocalisation analysis to identify genes likely associated with pneumonia. fastBAT analysis revealed eighteen genes, in chromosomes 9,15 and 16, to be associated with pneumonia risk (**Supplementary Table 2**). Sensitivity gene-based tests suggested the association of genes in chromosome 15, but not those in chromosomes 9 and 16, to be mediated by smoking (**Figure 2a**). Two genes, *HYKK* and *PBX3*, showed evidence of colocalisation in lung tissue (**Table 3**), but not whole blood. *EIF3C* showed suggestive evidence of colocalisation in the lung, and strong evidence of colocalisation in whole blood (**Supplementary Table 3**). While *IL27, CHRNA3* and *CHRNA5* have eQTL signals in the vicinity of pneumonia hits, our analysis suggests that the relationship between their expression and pneumonia is better explained by two neighboring independent causal variants (**Figure 2b**).

**Table 3.**
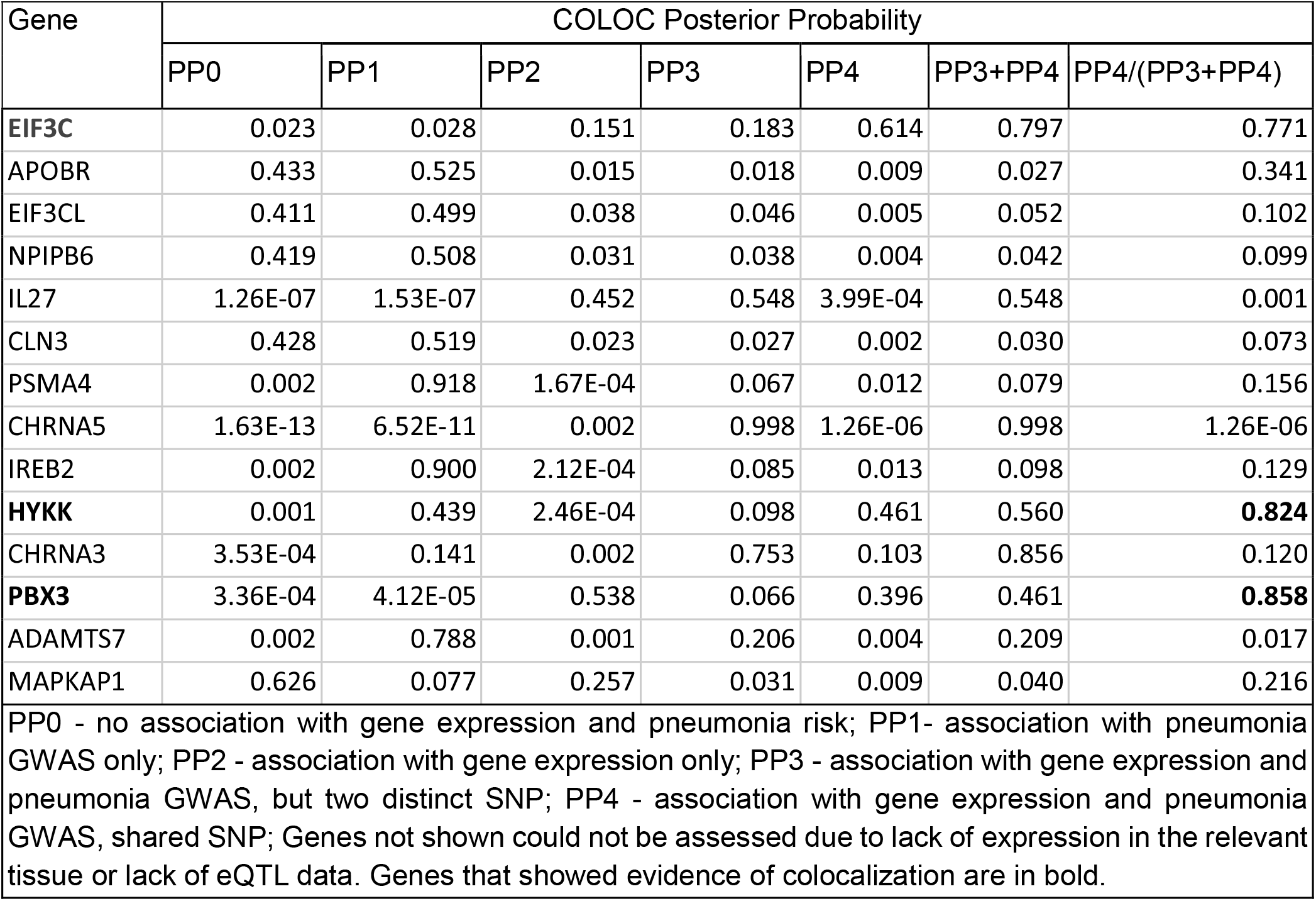
Colocalization of lung eQTLs with pneumonia GWAS loci.

**Figure 2.**
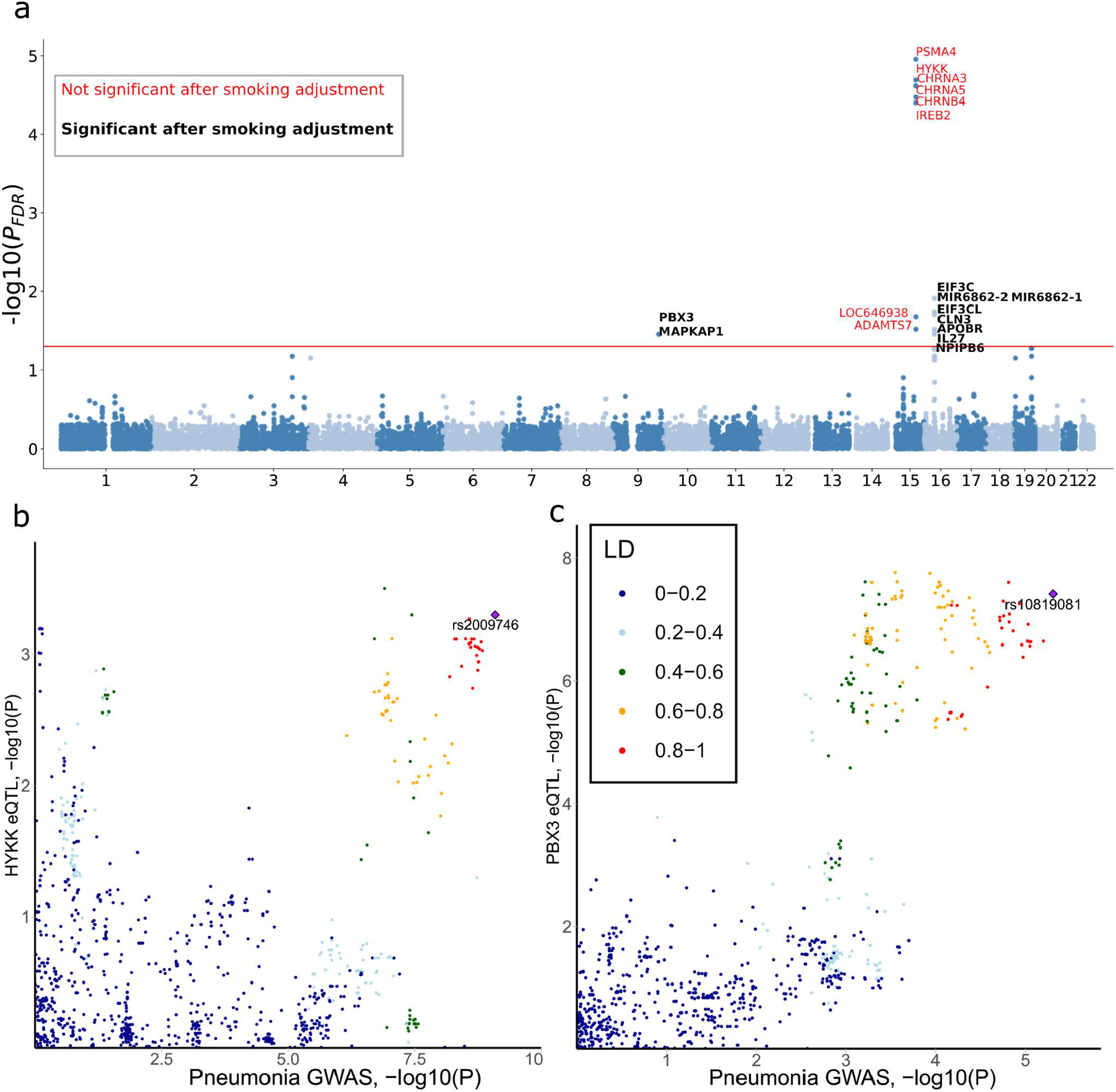
HYKK (CHR 15) and PBX3 (CHR9) eQTLs colocalise with pneumonia. a) Gene based test association results. Each dot represents a gene and its position on the y-axis corresponds to the p-value for association with pneumonia adjusted for multiple testing. Genes in bold (black) were robust to adjustment for smoking phenotypes, whereas genes in non-bold (red) font were not. Genes above the red line are significantly associated with pneumonia, and were assessed for eQTL colocalisation. b and c show colocalisation plots assessing shared signals between lung eQTLs and the pneumonia meta-analysis. Each dot represents a genetic variant. The × axis represents the evidence for association (-log10pvalue) between that variant and pneumonia. The y-axis represents the evidence for association between that variant and expression of the gene of interest. Colocalisation happens when there is a high level of co-occurrence between GWAS signals and eQTL signals. Two independent signals driving each trait would show two signals along the x and y axis respectively. Results shown only for HYKK (a) and PBX3 (b) as these genes showed evidence of colocalisation.

### LD-score genetic correlations

Across 1,522 traits studied, 552 traits displayed a genetic overlap with pneumonia at FDR < 5%. Traits with the strongest evidence of a genetic correlation with pneumonia included chronic obstructive pulmonary disease (COPD), “Wheeze or whistling in the chest in last year”, blood clot in the leg and myocardial infarction (**Figure 3**). Lifestyle factors such as current smoking showed a positive genetic correlation with pneumonia, indicating that variants that increase smoking behaviour also increase pneumonia risk. Genetic correlation between alcohol intake and pneumonia was conflicting, as the variable “*Alcohol usually taken with meals*” and “*Alcohol drinker status: Current*” had a negative genetic correlation with pneumonia. In contrast, the variable “Alcohol drinker status: Previous” displayed a positive genetic correlation with pneumonia. Traits related to mood or psychiatric disorders (such as depression and irritability), lifestyle variables (such as cycling to work and educational attainment), and biomarkers (such as immune cell count and C Reactive Protein [CRP]), among others, also showed significant genetic correlations with pneumonia (**Figure 3**).

**Figure 3.**
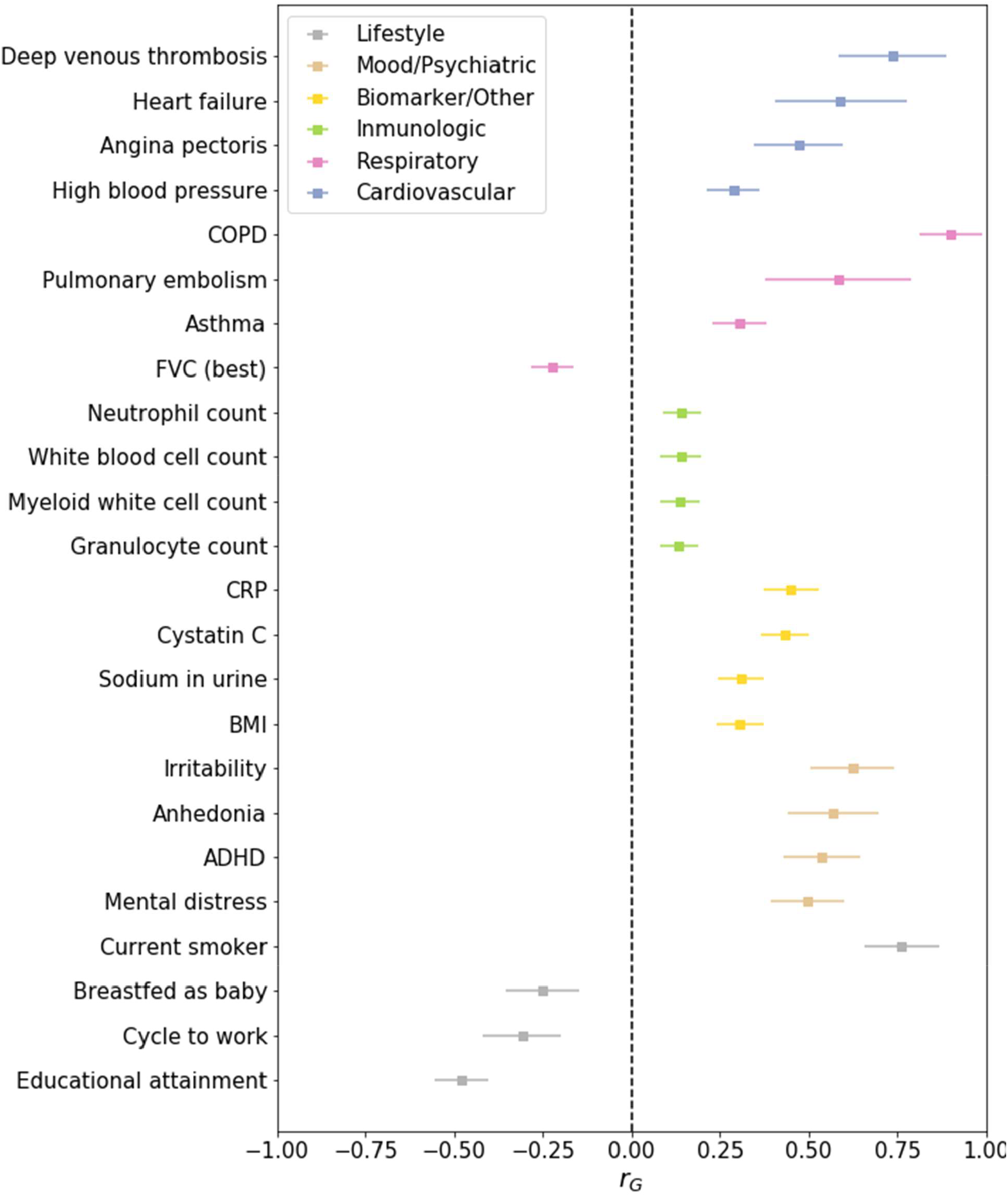
Pneumonia is genetically correlated with respiratory, circulatory, metabolic and lifestyle traits. Forest plot showing genetic correlations (rG) between pneumonia and traits of interest. Genetic correlations were estimated using bi-variate LD-score regression. All of the results shown are statistically significant. Due to space restrictions, the full results are available as Supplementary Data 1. Error bars represent standard errors of the genetic correlations.

### Genetic Causal Proportions

To assess whether the genetic correlations observed could be explained by a causal relationship, we performed a latent causal variable analysis. Forty four of the 552 traits with a significant (FDR < 5%) genetic overlap with pneumonia showed evidence of a causal association (**see methods**). LCV provided genetic evidence on several traits causally associated with pneumonia, including deep vein thrombosis (DVT), LDL (decreased), cholesterol (decreased) among other traits closely related to cardiovascular health, such as heart failure, arrhythmias and fibrillation. Evidence for DVT, hypertension, LDL and the cholesterol causal associations were further assessed using GSMR. This analysis showed a consistent result for DVT and hypertension, but no evidence of causality for LDL or cholesterol (**Supplementary Figure 2**). Traits highlighted as potential consequences of pneumonia included long-standing illness, lower forced vital capacity, anhedonia, pain, and taking omeprazole and co-codamol (**Figure 4** and **Supplementary Data 1**).

**Figure 4.**
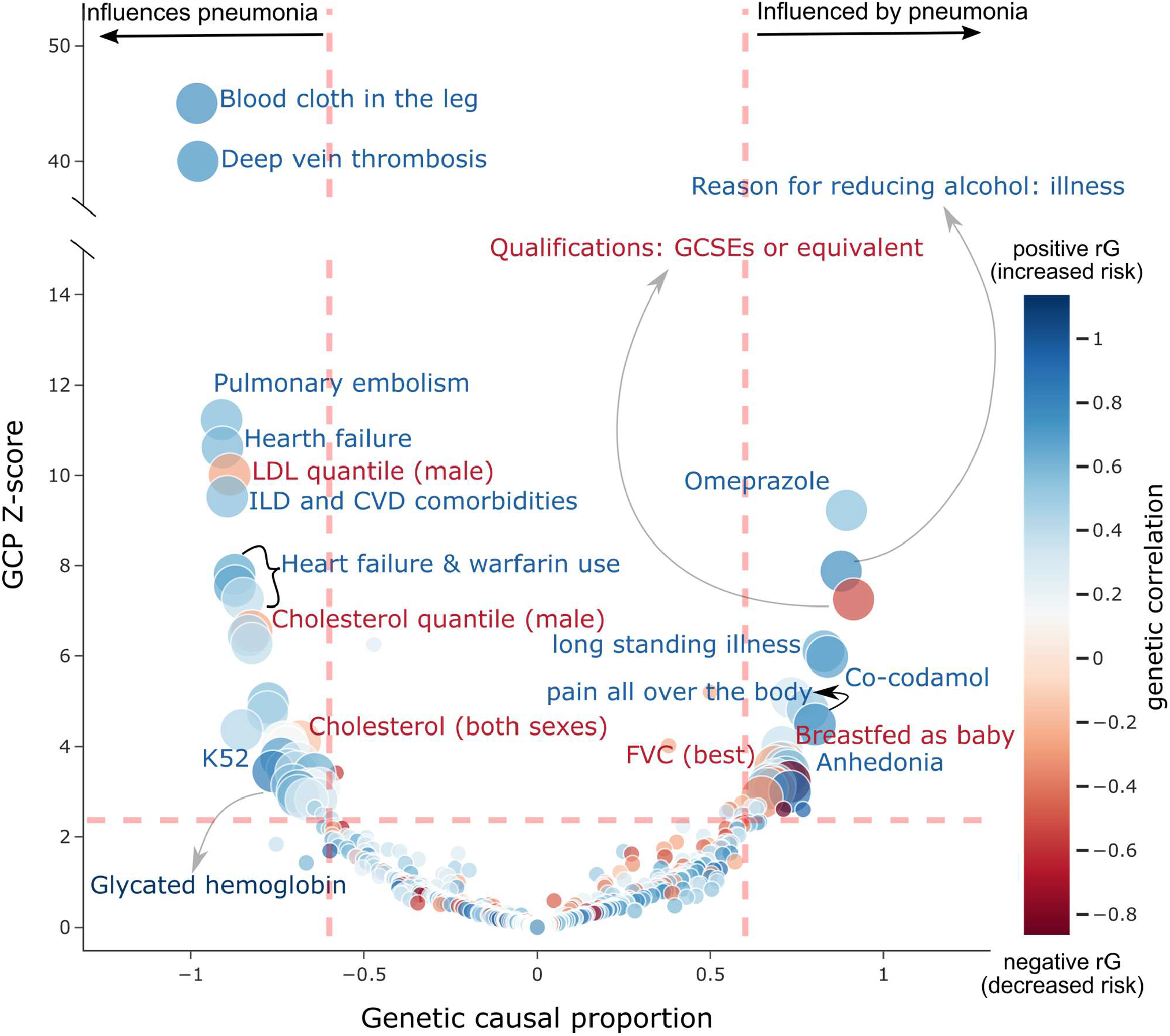
Pneumonia causal association analysis. Causal architecture plot showing the results of a phenome-wide latent causal variable analysis assessing the evidence for a causal association between pneumonia and other traits (see Methods). Each point represents a trait that showed a significant genetic correlation with pneumonia. The x-axis represents the genetic causal proportion; high values indicate evidence for a causal association between pneumonia and the trait of interest. Positive values indicate that pneumonia is likely to act as a risk factor for the trait (i.e. it *causes* the other trait). In contrast, negative values would highlight risk factors for pneumonia. Traits are coloured based on their genetic correlation with pneumonia and indicate the direction of the causal association (i.e. increasing risk or decreasing risk). Trait or trait category labels with a colour indicating the direction of the causal association have been added.

### Polygenic prediction of pneumonia

We performed polygenic prediction of pneumonia on the AGDS sample to assess the validity of our pneumonia GWAS. The prevalence of self-reported pneumonia history (∼2000 cases, ∼20%) in the AGDS sample was higher than pneumonia diagnosis in the UK Biobank (∼15k cases ∼3%) and FinnGen (∼10k cases ∼10%). Furthermore, the AGDS sample had a different age and sex composition from the UK Biobank (**Table 4)**. We assessed whether PRS derived from the pneumonia GWAS were associated with pneumonia in the AGDS cohort using a multivariate logistic regression (**see Methods**) and identified a statistically significant, but small in effect, association between pneumonia PRS and self-reported pneumonia OR=1.06 (95%CI=[1.01,1.12]; p=0.02) per standard deviation increase of pneumonia PRS.

**Table 4.** Target sample (AGDS) composition and demographics.

| **Table 4. Target sample (AGDS) composition and demographics** |  |  |
| --- | --- | --- |
|  | Cases | Controls |
| Sample size | 1206 (21%) | 4389 (78%) |
| Female (%)* | 919 (76%) | 3179 (72%) |
| Age (sd)* | 48.6 (14.6) | 42.5 (14.6) |
| Light smokers* | 955 (79%) | 3058 (69%) |
| Pneumonia PRS (sd)* | 0.14 (1.02) | 0.04 (0.98) |
| Data for unrelated participants of European ancestry used for the replication and PRS. *P<0.05 two sample t-test. |  |  |

### Sensitivity analyses

The genome-wide significant locus overlaps, and is in LD, with a set of well established smoking-associated variants including rs16969968.^20^ To assess whether the genetic associations for pneumonia are mediated by smoking, we performed several sensitivity analyses. A conditional association test showed that our top hit (rs2009746) evidence of association was reduced after adjusting for three independent smoking associated variants (p_rs2009746_=0.002; **Supplementary Table 4**). Nonetheless, an mtCOJO analysis suggested the associations between pneumonia and genes in chromosomes 16 and 9 to be independent from smoking (**Figure1** and **Figure2**). Finally, the association between pneumoniaPRS and self-reported pneumonia remained statistically significant after adjustment for smoking history both on the genetic and phenotypic level (**Supplementary Table 5**).

## DISCUSSION

Our findings highlighted eighteen genes, across chromosomes 6, 15 and 16 to be associated with pneumonia risk. We identified genes involved in general gene regulation (*PBX3, EIF3C*), iron regulation (*IREB2*), nicotine signaling (*CHRNA3/5*) and inflammatory processes (*IL27, APOBR*). Here, we integrated eQTL data with our GWAS results and performed colocalisation analysis to identify which genes have more robust evidence of association with pneumonia. Our analyses suggested *HYKK* and *PBX3* gene expression to colocalise with pneumonia. Notably, *PBX3* encodes a transcription factor whose deficiency has been linked to respiratory failure in mice.^22^ *HYKK* is an enzyme involved in lysine catabolism and was recently linked to nicotine metabolism.^23^ While our colocalisation analysis would suggest *HYKK, PBX3* and potentially *EIF3C* are associated with pneumonia through differential gene expression, other genes identified could be associated through mechanisms such as impairment or gain of function.

Genetic variants in 15q25.1 have been extensively linked with smoking.^24^ This complex region has also been previously associated with COPD^25^ and lung cancer^26^, and contains several compelling genes associated with nicotine addiction (*CHRNA3, CHRNB4, CHRNA5, HYKK*) and iron regulation (*IREB2*). We performed a sensitivity analysis and showed that 15q25.1 was not associated with pneumonia after adjusting for smoking history and cigarettes per day. Nonetheless, genes in other regions remained associated with pneumonia after adjusting for smoking. This is consistent with the observed high genetic correlation between the smoking-adjusted and unadjusted summary statistics. Moreover, polygenic prediction was also robust to adjustment for smoking history. Future efforts could leverage analyses such as pairwise GWAS or genomicSEM to further deconvolute the effects of smoking and respiratory disease. We consider this beyond the scope of the present study.

We discovered genetic correlations between pneumonia and biomarkers such as immune cell counts, cystatin C and sodium in urine. Consistently, Cystatin C and CRP levels have been linked to community-acquired pneumonia (CAP).^27,28^ Furthermore, lifestyle factors such as smoking, and lower socioeconomic status (as measured by the Townsend deprivation index) were genetically correlated with pneumonia. Finally, traits requiring healthy respiratory function such as *cycling to work* and *maximum workload during a fitness test* displayed a negative genetic correlation with pneumonia.

A genetic correlation between two traits could reflect causality between traits, or horizontal pleiotropy (genes acting on both traits independently of each other). Here, we performed LCV analyses to identify traits causally associated with pneumonia. Our results suggest that deep vein thrombosis (DVT) may causally increase risk of pneumonia. This result was further confirmed using GSMR. Previous studies have noted an association between these two diseases.^29^ Most studies suggest or assume that pneumonia causes DVT due to immobilization, hypoxia and inflammation. Hypoxia is one of the strongest predictors of pneumonia^29^ and has been shown to increase the incidence of thrombosis through the downregulation of protein S, a natural anticoagulant.^30^ Furthermore, tissue factor, along with coagulation related pathways, are known to be upregulated upon inflammation.^31^ Future studies should focus on further understanding of the intricate relationship between cardiovascular and respiratory diseases.

LCV also highlighted the involvement of cholesterol levels and LDL in decreasing the risk for pneumonia. Nonetheless, these results did not replicate in our GSMR analyses. Cholesterol is essential for cellular integrity and metabolism, and its dysregulation has been linked to a variety of diseases, including cardiovascular and pulmonary disease.^32^ Previous studies show that LDL and HDL trafficking influences multiple cell types in the lung.^33^ Class A scavenger receptors on alveolar macrophages uptake HDL as a source of vitamin E,^34^ which is an antioxidant that plays an essential role in the clearance of oxidized lipids that would otherwise result in cytotoxic and pro-inflammatory responses.^35^ Furthermore, cholesterol plays an essential role in protecting and covering the alveoli which prevents several pathological conditions.^36^ Thus, total cholesterol might protect from developing pneumonia through the relationship between cholesterol and immune homeostasis in the lung. Nevertheless, low levels of LDL have been associated with better lung function,^37^ and low HDL levels have been proposed as a poor prognosis marker for CAP.^38^ Moreover, a recent proteomic study in patients with sepsis secondary to pneumonia were found to have an impairment in lipid metabolism (lower total cholesterol, LDL cholesterol, as well as major apolipoprotein of LDL, ApoB).^39^ This is consistent with our gene based tests identifying the ApoB receptor (*APOBR*) as a potential pneumonia risk mediating gene. Overall our findings and the literature suggest that a dyslipidemic state, rather than specific levels of LDL influence pneumonia risk.

Some limitations of the present study must be acknowledged. We excluded participants of non-European ancestry to avoid biases due to population stratification. This limits the generalisability of our findings to populations of non-European ancestry. Furthermore, our results suggest that the genetic risk for pneumonia is highly complex, and several variants remain to be identified by more powered studies. Further evidence of this is the low polygenic prediction in an independent sample, which is still far from other traits where clinical relevance is starting to be considered. We replicated LCV findings using GSMR. Nonetheless, we could not attempt to replicate any of the causal associations where pneumonia was the exposure because our pneumonia GWAS was underpowered to be accurately used as an exposure. Finally, experimental approaches along with powered analyses considering not only smoking history but also smoking exposure and quantitative smoking measures are needed to claim, beyond any doubt, 15q25.1 to be associated with pneumonia over and above smoking.

In summary, pneumonia GWAS meta-analysis identified a region in 15q25.1 which has been previously linked to smoking, lung cancer and COPD. Gene-based tests association identified eighteen genes implicated in pneumonia risk in chromosomes 9, 15 and 16. Sensitivity analyses suggested the locus in chromosome 15 to be driven by smoking, but other associations were robust to adjustment for smoking related traits. eQTL colocalisation analysis in lung tissue suggested *HYKK, PBX3* and potentially *EIF3C* expression to colocalise with pneumonia. We identified traits with a significant genetic correlation and highlighted potential causally associated traits, including DVT and lipid homeostasis. Finally, validation of our GWAS was obtained by polygenic prediction of self-reported history of pneumonia in an independent sample. Polygenic prediction was robust to adjustment for smoking history, suggesting some independence of our GWAS signals from smoking history. Increasing statistical power could help identify additional genetic targets which will, in turn, enable the development of new therapeutics and patient risk stratification based on genetic risk.

## Data Availability

The GWAS summary statistics and results for the GWAS downstream analyses will be made available after peer-review. Code used for this study and results are available upon request.

## ACKNOWLEDGMENTS

This research was conducted using data from the UK Biobank resource under application number 25331. We want to acknowledge the participants and investigators of the FinnGen study. Data collection for the Australian Genetics of Depression Study was possible, thanks to funding from the Australian National Health & Medical Research Council (NHMRC) to N.G.M. (GNT1086683). A.I.C. and K.X.V.P. are both supported by UQ Research Training Scholarships from The University of Queensland (UQ). P.F.K. is supported by an Australian Government Research Training Program Scholarship from Queensland University of Technology (QUT). M.E.R. thanks support of the NHMRC and Australian Research Council (GNT1102821).

## AUTHOR CONTRIBUTIONS

AIC conceived the study. AIC and PFK performed the analyses with aid and input from NGM, GCP and MER. KXVP and LGM helped interpreting the results. NGM designed and directed the AGDS study. All authors collaboratively wrote the manuscript.

## DATA AVAILABILITY

The full GWAS summary statistics for this study will be made available through the NHGRI-EBI GWAS Catalogue (https://www.ebi.ac.uk/gwas/downloads/summary-statistics) upon publication. Individual level data for UK Biobank participants are available to eligible researchers through the UK Biobank (www.biobank.ac.uk). Results for the GWAS downstream analyses have been made available in CTG-VIEW (https://view.genoma.io). Code used for this study is available upon request.

